# Voice-Enabled Response Analysis Agent (VERAA): Leveraging Large Language Models to Map Voice Responses in SDoH Survey

**DOI:** 10.1101/2023.09.25.23295917

**Authors:** Rishivardhan Krishnamoorthy, Vishal Nagarajan, Hayden Pour, Supreeth P. Shashikumar, Aaron Boussina, Emilia Farcas, Shamim Nemati, Christopher S. Josef

**Affiliations:** University of California San Diego Health, Department of Biomedical Informatics, San Diego, CA; Healcisio, Inc., La Jolla, CA; Qualcomm Institute, the San Diego division of the California Institute for Telecommunications and Information Technology (Calit2)

## Abstract

Social Determinants of Health (SDoH) have been shown to have profound impacts on health-related outcomes, yet this data suffers from high rates of missingness in electronic health records (EHR). Moreover, limited English proficiency in the United States can be a barrier to communication with health care providers. In this study, we have designed a multilingual conversational agent capable of conducting SDoH surveys for use in healthcare environments. The agent asks questions in the patient’s native language, translates responses into English, and subsequently maps these responses via a large language model (LLM) to structured options in a SDoH survey. This tool can be extended to a variety of survey instruments in either hospital or home settings, enabling the extraction of structured insights from free-text answers. The proposed approach heralds a shift towards more inclusive and insightful data collection, marking a significant stride in SDoH data enrichment for optimizing health outcome predictions and interventions.

## Introduction

Over the past decade, health systems and providers have invested significant efforts to simultaneously reduce burgeoning costs while improving the quality of care provided. Recent research has demonstrated that Social Determinants of Health (SDoH) contributed to 40-50% of the cost structure in Medicare and Medicaid (1) and served as the most important contributor towards health outcomes (2). SDoH encompass the societal and economic conditions influencing health. Variables such as education, socioeconomic status, neighborhood environment, employment, and social support networks have begun to be viewed not just as background factors, but as critical elements that can steer the course of individual health trajectories. Regrettably, structured SDoH for planning or predictive modeling is not always available (3). A recent survey found that SDoH data elements can be missing 20-89% of the time (4). Additionally, new reporting requirements stemming from the Center for Medicare and Medicaid Services (CMS) Framework for Health Equity have mandated that health systems begin collecting SDoH data in 2024.

Traditional approaches to collection of health survey data generally require a one-to-one interview conducted as part of the intake or admission process, which can increase system workload and is limited by the languages spoken by the patient and interviewer. Limited English proficiency (LEP) in the United States can be a barrier to accessing health care services and understanding health (5). While most providers use bilingual staff or interpreters to communicate with LEP patients, challenges such as availability, quality, and cost of these services limit their effectiveness. Furthermore, as patient surveillance extends into the home environment (6) the collection of accurate, up-to-date information presents additional challenges since the availability of a broadband internet connection or a human interviewer can be limited. Clinicians and researchers are thereby challenged to develop collection modalities that not only capture a wide breadth of information but also resonate with the lived experiences of a heterogeneous populace.

A recent study has established a strong association between LEP and sepsis mortality (7). While the causal mechanism is unknown, LEP is known to be associated with greater difficulties in accessing medical care, and language barriers can impede providers’ ability to take an appropriate clinical history that may lead to clinical errors or delays in care. In our previous work, we have demonstrated that SDoH (8) and wearable (9,10) data can dramatically improve the accuracy of sepsis readmission scores. However, SDoH factors are often poorly captured in electronic health records (EHRs) and are not available at the time of hospital discharge. As such, there is an unmet need for patient-centered communication technologies that can facilitate multilingual health data collection in diverse settings including inpatient and outpatient healthcare facilities, as well as patients’ homes (11). Voice assistants provide novel opportunities for data collection as well as addressing some of the barriers that patients encounter daily while managing their health (12).

Home implementation requires a device capable of 1) periodically accessing the EHR in a HIPAA-compliant manner, 2) integrating wearable (e.g., Fitbit) data, and 3) gathering structured and unstructured responses from patients. To achieve this we have built the My Companion Pet (MCoPet) device as described in Figure 1. The bedside, stand-alone MCoPet utilizes a Raspberry Pi board in association with a speaker and microphone, and seamlessly integrates with an EHR and wearable sensors. MCoPet is capable of bi-directional EHR communication via the HL7 FHIR (Fast Healthcare Interoperability Resources) APIs, retrieving wearable information from Fitbit via the Fitbit Web API, and locally running predictive algorithms. The most distinguishing feature of MCoPet has been the incorporation of a large language model (LLM) (i.e., Llama-2 (13)) in conjunction with a multilingual, voice-to-text transcription system (via the Whisper (14) automatic speech recognition deep learning model). This system is capable of transcribing the free responses to survey questions and mapping them to specific survey answers (e.g., multiple choice options) in a “conversational” manner. Owing to the recent development and release of publicly-available LLMs, there has been little to no evidence generated to support the use of LLMs for mapping free responses to structured survey responses. Given the potential for transformative change, investigating the application of LLMs for use in collecting SDoH information is not just a matter of improving understanding and predictive model performance, but a collaborative journey aimed at addressing the complex interplay of societal dynamics and individual health narratives.

**Figure 1.**
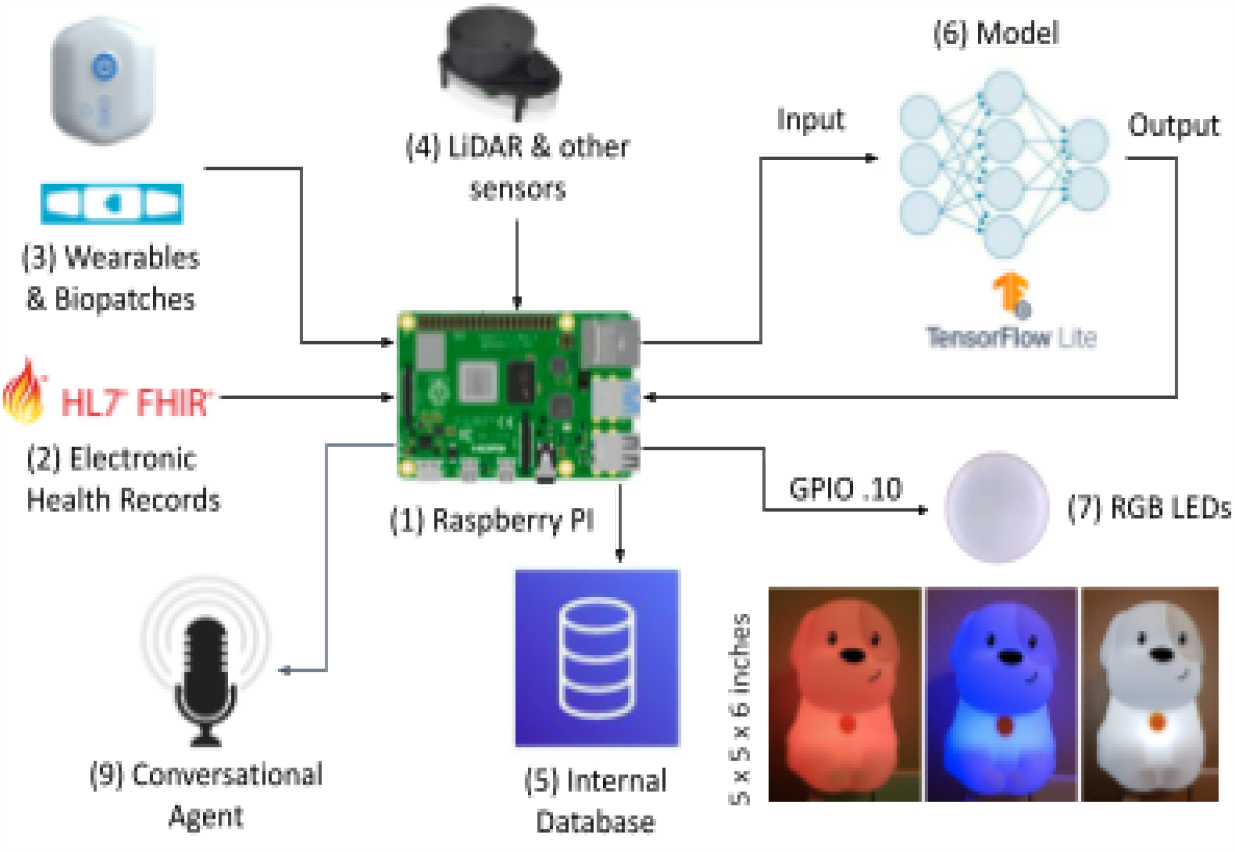
Schematic diagram of the My Companion Pet (MCoPet) device: A Raspberry PI (1) is used to communicate with external data sources such as EHR (2), wearables and biopatch sensors (3), and other attached sensors such as LIDAR (4). Data is harmonized in an internal database (5). Predictive risk scores are locally calculated using Python and TensorFlow Lite models (6) The resiling dichotomized risk scores (above or bekw threshold) are used to control two or more LED light sources (7), to ndicate risk for sepsis, sepbc shock, or deterioration, among others. A conversational agent (9) conducts surveys (See Figure 2).

## Methods

We have previously conducted predictive modeling work addressing sepsis readmission (9,10) using SDoH survey data (8) from the All Of Us dataset (15). In this previous work, we identified a subset of 28 questions across four surveys that significantly improved readmission prediction accuracy for sepsis. This subset of questions (see the Results section) were derived from the All of Us “The Basics Survey” (B), “The Lifestyle Survey” (L), “The Healthcare Access and Utilization Survey” (H), and “The Overall Health Survey” (O).

For each of the 28 questions, we generated 10 distinct scripted replies encompassing all answer choices (i.e, a total of 280 potential responses). A sample of these scripted replies is provided in Table 1 which are mapped to survey answers contained in Table 2. A panel of two English speakers and one Spanish speaker then read the generated scripts for a total of 20 English audio responses and 10 Spanish audio responses per question. This amounted to a total of 840 distinct audio clips. The processing pipeline is described in Figure 2. The survey begins with a question from the device which the respondent then answers. The respondent’s audio is then converted into text using voice-to-text transcription via the Whisper automatic speech recognition deep learning model. The transcribed text is then passed to Llama-2 with instructions to map the text to the most appropriate survey response. Each of the 280 scripted responses were examined by a panel of three reviewers who through consensus mapped the scripted response to one of the structured survey answers. This “ground truth” was then used for evaluating the LLM’s mapping accuracy.

**Figure 2.**
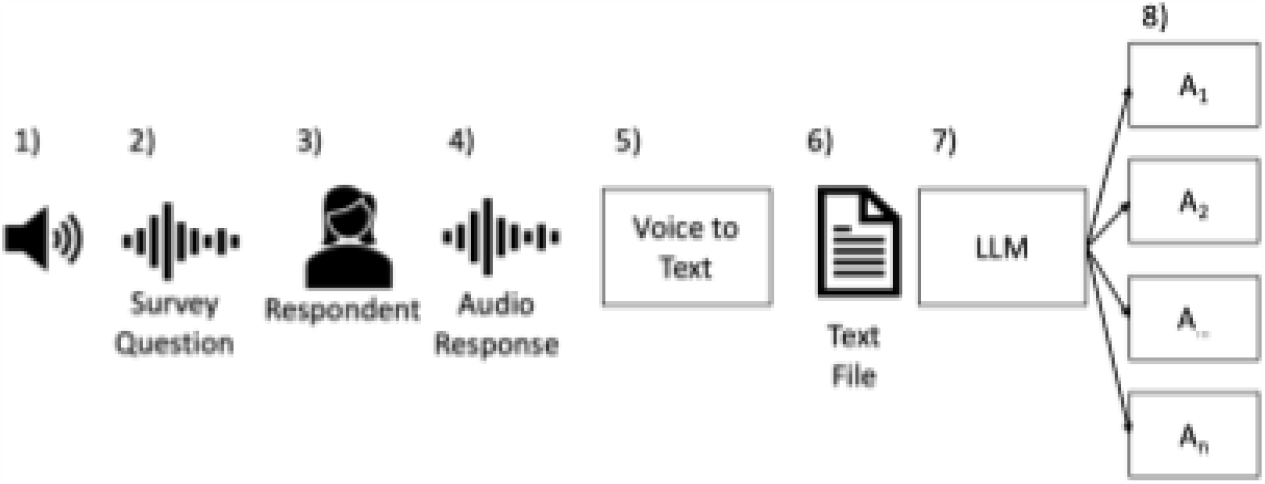
MCoPct Conversational Agent workflow: 1) MCoPct poses a 2) survey question to the 3) respondent who generates an 4) audio response (see sample audio response in Table 1), which is translated into English text via 5) Whisper. This 6) text file is then passed to rhe 7) L.L.M with instructions to map the text to one of the survey question 8) answers (see Table 2).

**Table 1:** Sample of Free Text Responses in English and Spanish.

| Table 1: Sample of Free Text Responses in English and Spanish |  |
| --- | --- |
| **B8.** Do you own or rent the place where you live? | **B8.** Spanish Translation of question B8 removed per MEDRXIV policy |
| 1. "I own my home; bought it a few years back."<br>2. "We rent a cozy apartment in the city center."<br>3. "I'm in another arrangement; I live with my parents for now."<br>4. "We own a small house on the outskirts of town."<br>5. "Currently, I rent a studio near my workplace."<br>6. "It's another arrangement; I'm staying with a friend temporarily."<br>7. "I own a condominium downtown, which I purchased last year."<br>8. "Renting for now, but hoping to buy in the next couple of years."<br>9. "I'm in another arrangement; it's a housing cooperative, so it's a bit different."<br>10. "I rent a townhouse with a couple of roommates." | 1. Translated response removed per MEDRXIV policy.<br>2. Translated response removed per MEDRXIV policy.<br>3. Translated response removed per MEDRXIV policy.<br>4. Translated response removed per MEDRXIV policy.<br>5. Translated response removed per MEDRXIV policy.<br>6. Translated response removed per MEDRXIV policy.<br>7. Translated response removed per MEDRXIV policy.<br>8. Translated response removed per MEDRXIV policy.<br>9. Translated response removed per MEDRXIV policy.<br>10. Translated response removed per MEDRXIV policy. |

**Table 2:** Answer Choices for SDoH Survey Questions.

| Table 2: Answer Choices for SDoH Survey Questions |  |  |  |  |
| --- | --- | --- | --- | --- |
| B1 | B2, L1-5 | B3 | B4 | B5 |
| Less than \$10,000<br>\$10,000- \$24,999<br>\$25,000-\$34,999<br>\$35,000-\$49,999<br>\$50,000- \$74,999<br>\$75,000-\$99,999<br>\$100,000- \$149,999<br>\$150,000- \$199,999<br>\$200,000 or more<br>Prefer not to answer | Yes<br>No<br>Don't know<br>Prefer not to answer | Employed for wages (This can be part- time or full-time)<br>Self-employed<br>Out of work for 1 year or more<br>Out of work for less than 1 year<br>A homemaker<br>A student<br>Retired<br>Unable to work (disabled)<br>Prefer not to answer | Never attended school or only attended kindergarten<br>Grades 1 through 4 (Primary)<br>Grades 5 through 8 (Middle school)<br>Grades 9 through 11 (Some high school)<br>Grade 12 or GED (High school graduate)<br>1 to 3 years after high school (Some college, Associate's degree, or technical school)<br>College 4 years or more (College graduate)<br>Advanced degree (Master's, Doctorate, etc.)<br>Prefer not to answer | Yes<br>No<br>Prefer not to answer |
| B6 | H1-12 | O1 | O2 | O3-5 |
| Own<br>Rent<br>Other arrangement | Yes<br>No<br>Don't know | Extremely<br>Quite a bit<br>Somewhat<br>A little bit<br>Not at all | Never<br>Rarely<br>Sometimes<br>Often<br>Always | Excellent<br>Very good<br>Good<br>Fair<br>Poor |

While our conversational agent has been purposely built for edge-computing devices, we leveraged cloud computing resources to speed prototyping and optimization of the conversational agent. We utilized a HIPAA-compliant AWS instance of type g5.4xlarge, comprising an NVIDIA A10 GPU coupled with 24GB VRAM and 64GB memory running Windows Server 2022. The 3-bit quantized version of the Llama 2 70-Billion parameter variant was employed where a total of 44 layers of the model were offloaded to GPU for accelerated performance. The pure C++ implementation of the Llama-2 model, called Llama.cpp, was used for inference resulting in generating 1.8 tokens per second using the AWS instance’s resources.

We performed three experiments: MCoPet Hybrid English, Cloud English, and Cloud Spanish. In the first experiment, a hybrid pipeline was set up where the English audio was transcribed to text using Whisper’s base model (74 Million parameters) on the MCoPet device, and text files were then passed to Llama-2 70B hosted on AWS. In the second and third experiments, the English and Spanish audio files were passed to the AWS instance where Whisper’s large model (1.550 Billion parameters) was used to produce audio transcription that Llama-2 70B used for inference. Furthermore, in an effort to localize the computation pipeline to the MCoPet device, we ported the 7B parameter variant of the Llama-2 model onto the MCoPet module and observed ∼300 seconds or 5 minutes to process one response. In comparison, the Llama-2 70B parameter model on the AWS instance processed each response at an average rate of 1 minute and also generated accurate mappings better than the Llama-2 7B model. Therefore, for the purpose of this paper, we focused on the full Llama-2 70B model on AWS, and the Hybrid English experiments were limited to just using Whisper on the MCoPet device.

### Results

The performance of MCoPet Hybrid English, Cloud English, and Cloud Spanish conversational agents are contained within Table 3. The accuracy (ACC) for a question represents the accuracy of matching 20 English responses and 10 Spanish responses to the set of response choices for each question.

**Table 3:** LLM Survey Mapping Accuracy by Question.

| Table 3: LLM Survey Mapping Accuracy by Question |  |  |  |  |
| --- | --- | --- | --- | --- |
| Question |  | Hybrid English (%ACC) | Cloud English (%ACC) | Cloud Spanish (%ACC) |
| B1 | What is your approximate annual household income from all sources? | 75 | 75 | 70 |
| B2 | Are you covered by health insurance or some other kind of health care plan? | 100 | 100 | 100 |
| B3 | What is your current employment status? Please select 1 or more of these categories. | 90 | 100 | 100 |
| B4 | What is the highest grade or year of school you completed? | 90 | 90 | 100 |
| B5 | In the past 6 months, have you been worried or concerned about NOT having a place to live? | 95 | 100 | 100 |
| B6 | Do you own or rent the place where you live? | 100 | 100 | 100 |
| L1 | Have you smoked at least 100 cigarettes in your entire life? (There are 20 cigarettes in a pack)? | 100 | 100 | 100 |
| L2 | Have you ever used an electronic nicotine product, even one or two times? (Electronic nicotine products include e- cigarettes, vape pens, hookah pens, personal vaporizers and mods, e-cigars, e-pipes, and e-hookahs.) | 95 | 100 | 100 |
| L3 | Have you ever smoked a traditional cigar, cigarillo, or filtered cigar, even one of two puffs? | 100 | 100 | 100 |
| L4 | Have you ever smoked tobacco in a hookah, even one or two puffs? | 95 | 100 | 100 |
| L5 | Have you ever used smokeless tobacco products, even one or two times? (Smokeless tobacco products include snus pouches, Skoal Bandits, loose snus, moist snuff, dip, spit, and chewing tobacco.) | 95 | 100 | 100 |
| During the past 12 months, were any of the following scenarios true? |  |  |  |  |
| H1 | You saw or talked to a general doctors who treated a variety of illnesses ( a physician in general practice, primary care, family medicine, or internal medicine) | 90 | 100 | 100 |
| H2 | You used alternative therapies to save money. | 85 | 90 | 90 |
| H3 | You skipped medication doses to save money. | 80 | 100 | 90 |
| There are many reasons people delay getting medical care. Have you delayed getting care for any of the following reasons in the PAST 12 MONTHS? |  |  |  |  |
| H4 | Could not afford the copay. | 85 | 100 | 90 |
| H5 | Your deductible was too high/or could not afford the deductible. | 80 | 90 | 80 |
| H6 | You had to pay out of pocket for some or all of the procedure. | 80 | 100 | 90 |
| H7 | Did not have transportation. | 80 | 80 | 80 |
| H8 | You provide care to an adult and could not leave him/her. | 75 | 80 | 80 |
| H9 | You could not get child care. | 80 | 90 | 80 |
| H10 | You live in a rural area where distance to the health care provider is too far. | 75 | 90 | 80 |
| DURING THE PAST 12 MONTHS, was there any time when you needed any of the following, but didn't get it because you couldn't afford it? |  |  |  |  |
| H11 | To see a specialist. | 80 | 80 | 80 |
| H12 | Follow-up care. | 90 | 90 | 90 |
| O1 | How confident are you in filling out medical forms by yourself? | 95 | 95 | 80 |
| O2 | How often do you have someone help you read health-related material? | 70 | 100 | 80 |
| O3 | In general, how would you rate your physical health? | 80 | 95 | 80 |
| O4 | In general, would you say your quality of life is? | 75 | 95 | 90 |
| O5 | In general, how would you rate your satisfaction with your social activities and relationships? | 85 | 100 | 80 |

A boxplot of the performance of our proposed system across the MCoPet Hybrid English approach, Cloud-based English, and Cloud-based Spanish approach is shown in Figure 3. The median and interquartile accuracy for the MCoPet Hybrid English approach was 85% [80%-95%]. Similarly, the accuracy for the Cloud English and Cloud Spanish approaches was 100% [90%-100%] and 90% [80%-100%], respectively. Since all the approaches use the same LLM (i.e., Llama-2) in the backend for mapping, the variation in performance between the Hybrid and Cloud English approaches can be attributed to Whisper. While the cloud based approaches support Whisper’s large model, MCoPet can only accommodate Whisper’s base model. The difference between performance of the Whisper base model and large model is prominently showcased in Figure 3.

**Figure 3.**
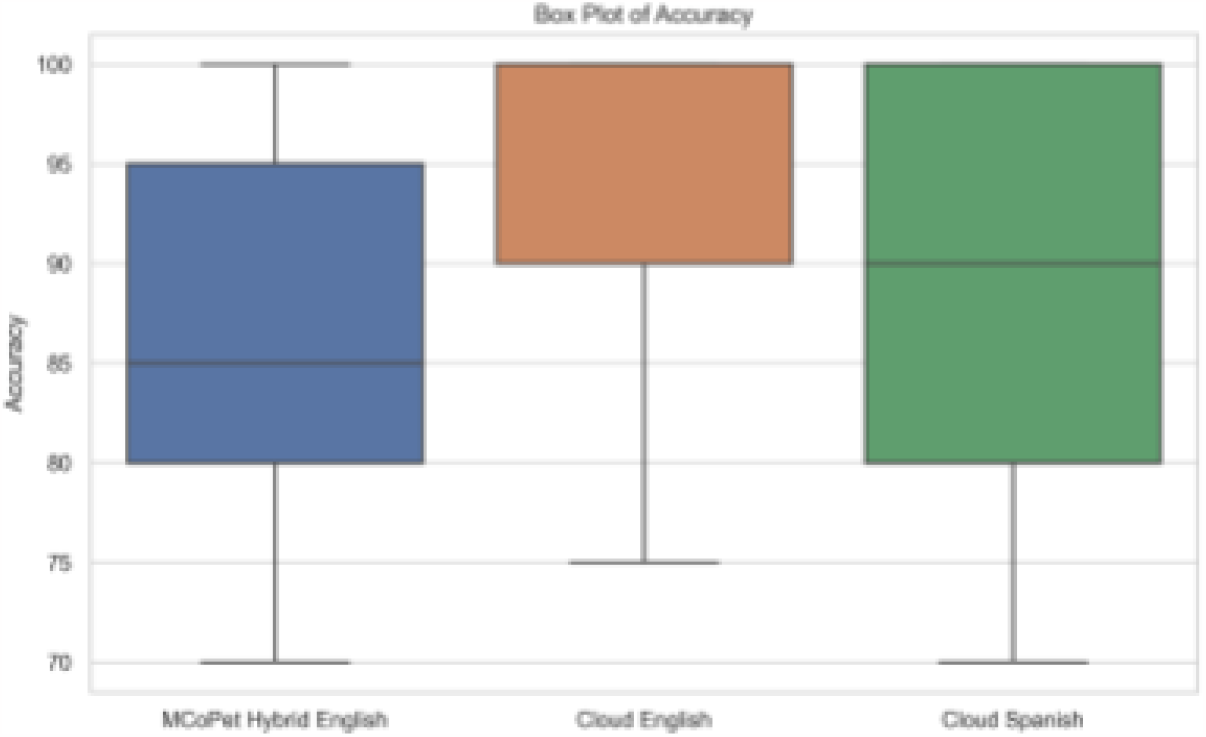
Boxplot delineating interquartile ranges of accuracies of MCoPet Hybrid English, Cloud English, Cloud Spanish approach.

It can be observed that the performance of Llama-2 is discernibly lower when the questions involve mathematical comparisons (e.g., question B1). For example, the task in question B1 is for the LLM to match the patient’s response to the correct bin of annual household income range. Llama-2 can correctly match the response when the annual income is equal to the lower or upper bound of the range, but not when it is within the range. This suggests that Llama-2 lacks the ability to perform relational operations on numerical values. This limitation is consistent with the findings of Imani et al., who showed that LLMs can generate incorrect answers with certain formulations of math questions (16).

Furthermore, our experiments revealed consistent mispredictions in scenario-based question responses (e.g., H question series). These mispredictions often involved responses containing the phrase “Don’t know”, which, upon closer examination, actually contained information indicative of a “Yes” or “No” prediction. For instance, consider the question, “Did you delay getting medical care because of transportation?” and a patient’s response of “Don’t know if it was solely due to transportation, but there were times I couldn’t make it.” Despite the response implying a delay in medical care, Llama-2 focused on the “Don’t know” aspect and incorrectly predicted “Don’t know” instead of “Yes”. This issue may be attributed to the enforced Llama-2’s deterministic nature, which is to ensure consistency and accuracy across all question-response pairs.

We observed that the transcription by Whisper’s base model was inaccurate in certain instances, sometimes omitting spoken words at the end of sentences, which led to reduced accuracy for the Hybrid English compared to Cloud English approach. Similarly, we noticed that Whisper’s large model on the cloud sometimes failed to accurately translate words from Spanish to English. For instance, the words “Rarely” and “Fair” that were answered in Spanish for questions O2 and O3 were incorrectly transcribed and translated to “Almost never” and “Just” leading to mispredictions by the LLM. These factors contributed to reducing the accuracy of the Cloud Spanish approach as opposed to the Cloud English approach.

## Discussion

Previous work by Nguyen et al. has identified that most prior work in the development of conversational agents has suffered from the limitations of constrained input, namely that only certain responses can be accepted (17). The primary finding of this proof of concept study is that LLMs can be utilized to map unconstrained, free responses to discrete survey answers with relatively high accuracy across more than one language, which demonstrates a move towards unconstrained modalities of information collection. The incorporation of an LLM based conversational agent within MCoPet presents an opportunity to conduct “context aware” interactions that account for additional sensors and EHR data.

Improvements in communication between patients and their care teams, especially for LEP patients are likely to contribute to more timely and equitable care. Conversational agents like the one proposed in this study offer a pathway for removing some of the barriers imposed by language by offering a location agnostic (e.g., home, hospital, etc.), low cost method for capturing accurate, complex information from patients without the presence of an interviewer. A successful implementation of such a tool could rapidly expand the amount of health-related information collected without a concomitant increase in labor. While the focus of this work was limited to producing responses to an existing structured survey instrument, the findings are extensible to the collection and interpretation of multilingual free responses into structured data. Use cases include but are not limited to: soliciting symptoms, gauging responses to interventions or medications, patient satisfaction, among others. Currently, surveys issued at scale often omit free responses owing to the time and labor necessary to convert the responses into some form of usable information; however, applications of LLM’s greatly reduce if not eliminate these limitations. Future surveys may now be able to avoid the use of structured responses which oftentimes don’t afford respondents the option of qualifying or explaining their answers.

We acknowledge that this investigation has certain limitations. In some circumstances a tablet based survey may afford users a shorter survey duration; however, this approach does not lend itself well to the collection and analysis of unstructured, free responses and is not as easily adaptable to populations with language or literacy barriers. The questions utilized only represent a subset of a larger SDoH survey set curated and validated by the All of Us Data consortium. The inclusion of additional questions may uncover problematic questions that are not appropriately mapped by a LLM. Additionally, the conducted experiments were limited to a set of pre-formed responses in order to compare accuracy across languages and speakers given the same response. The full spectrum of wild type responses still stands to be evaluated. The experiments were limited to two languages and do not fully represent the diverse range of cultural or lingual backgrounds treated within the US healthcare system; however, the method is currently extendable to 99 different languages and the number of languages and performance across languages is expected to rapidly improve.

Future development of this work will increase the number of responses and compare them to ground truth answers from traditionally administered surveys gathered as part of the aforementioned CMS reporting requirement set to begin in 2024. LLMs could also be applied to existing text in the medical record to prepopulate responses to survey questions. The respondent could then confirm that this information is correct or adjust accordingly during their interactions. Lastly, focus group testing with community health representatives would better elucidate technological literacy and cultural barriers that may lead to hesitancy or inaccuracies associated with verbal, health based surveys.

## Conclusion

This paper presents a novel approach to collect and analyze multilingual SDoH survey data using a voice-enabled conversational agent that leverages automatic speech recognition and large language models. VERAA transcribes and maps voice responses in multiple languages to structured survey answers with high accuracy. The proposed technology can be integrated with a low-cost, edge-computing device that can communicate with EHRs and wearable sensors, enabling data collection in diverse outpatient settings. The results of this proof-of-concept study demonstrate the potential of LLMs to facilitate more inclusive and insightful SDoH data collection, which can improve health outcome predictions and interventions. Future work will involve testing VERAA with real patients and expanding the scope of survey instruments and languages supported.

## Data Availability

All data produced in the present work are contained in the manuscript.

## Acknowledgements

S.N. is funded by the National Institutes of Health (R35GM143121). He is a co-founder of Healcisio Inc., which is focused on commercialization of advanced analytical decision support tools. Mr. Boussina is funded by the National Library of Medicine (#2T15LM011271-11). The opinions or assertions contained herein are the private ones of the author and are not to be construed as official or reflecting the views of the NIH or any other agency of the US Government. We would like to thank Drs. Robert El-Kareh and Dr. Robert Owens for their discussions regarding workflow implementation of the proposed conversational agent. Additionally, we would like to acknowledge Ised Gongora for help with Spanish translation and audio recordings.

## Data Sharing

The survey data used in this study, along with the evaluation codes and audio files, will be made available on GitHub for public access and reproducibility. The GitHub repository link will be provided in the final version of the paper.

## Paper Notes (Will Not be included in final manuscript)

### Program Track

Data/Science Artificial Intelligence

### Theme

Harnessing the Power of Large Language Models in Health Data Science

### Keywords (limit 3)

- Clinical decision support for translational/data science interventions
- **Clinical and research data collection, curation, preservation, or sharing**
- Ethical, Legal, and Social Issues
- Fairness and disparity research in health informatics
- Implementation Science and Deployment
- Knowledge representation, management, or engineering
- Learning healthcare system
- **Machine learning, Generative AI, and predictive modeling**
- Mobile Health, wearable devices and patient-generated health data
- Natural Language Processing
- Outcomes research, clinical epidemiology, population health
- Patient centered research and care
- **Social determinants of health**

### Titles

- “The Future of Health Surveys: Unveiling the Potential of LLMs in Analyzing Voice Transcribed SDoH Survey Data”
- “Improving SDoH Survey Outcomes with LLM-Based Voice Transcription Mapping: A Case Study”
- “A New Frontier in Health Equity: Leveraging LLMs for Enhanced SDoH Survey Analysis”
- “Realizing the Potential of LLMs in Facilitating Comprehensive SDoH Survey Analyses”
- “A Proof-of-Concept Study: LLM-Based Mapping of Voice Transcribed Answers to SDoH Surveys”
- “Pilot Study on the Efficacy of LLMs in Analyzing Voice Transcribed Responses to SDoH Surveys”
- “Exploring the Frontiers of AI in Healthcare: A Preliminary Study on LLM-Based SDoH Survey Analysis”
- “Voice to Insight: Leveraging Large Language Models for SDoH Survey Analysis”
- “Automated Mapping of Voice Transcribed Responses to SDoH Surveys: A Novel Approach”
- “Towards Intelligent SDoH Surveys: A Voice-Activated Response Mapping System”
- “Voice-Activated SDoH Survey Analysis through Large Language Model Integration”
- “Innovations in Public Health: Utilizing LLMs for SDoH Survey Voice Responses
- MUSA: Multilingual Survey Analysis using Automatic Speech Recognition and Large Language Models
- “Voice-Enabled Response Analysis Agent (VERAA): Leveraging Large Language Models to Map Voice Responses in SDoH Surveys”

### Learning Objective

Upon completion of this educational experience individuals should be able to select and use appropriate transcription software and large language models to map free text audio responses to answers fields on surveys.

